# Opioid distribution trends in California post recreational marijuana legalization

**DOI:** 10.1101/2021.02.20.21252025

**Authors:** Michelle N. Anyaehie, Christian Pardo, Elijah J Johnson, Chucks Anachebe, Brian J. Piper

## Abstract

The opioid epidemic has risen to an all-time high. Harm reduction and prevention policies have not alleviated this crisis. Recent investigations have highlighted the efficacy and safety of marijuana-based products for pain management. Providing alternative pain treatment options may help mitigate the opioid epidemic. The distribution of codeine, fentanyl, hydrocodone, morphine, and oxycodone per 100K people and by 3-digit zip codes and overdose rates from 2014 to 2018 in California, which legalized recreational marijuana in 2016, were compared to Texas, where marijuana is functionally prohibited. Drug weights were obtained from the Automation of Reports and Consolidated Orders System and converted to oral morphine milligram equivalents. Overdose data was retrieved from the Centers for Disease Control’s WONDER database. California (−43.7%) and Texas (−27.3%) showed significant reductions in cumulative opioid distribution from 2014 to 2018. Opioid distribution per 100K people decreased −38.9% in California relative to −26.4% in Texas. Opioid and heroin overdoses increased between 1999 and 2019 by +11.6% in California but +272.7% in Texas. This evidence supports marijuana legalization as a mitigating factor to the opioid epidemic. Continued studies on safer pain management alternatives and policies will help identify measures that help combat the opioid epidemic.

## Introduction

The opioid epidemic is characterized by a steep increase in opioid prescription rates followed by a rise in opioid-related deaths in the early 1990s. Opioid prescriptions quadrupled from 1998 to 2008 and resulted in alarming overdose rates (Jones et al., 2018). Nationwide opioid prescription rates steadily increased from 2006 to 2012 but started decreasing from 2013 to 2018 (CDC Injury Center, 2020), showing there has been an effort to alleviate the crisis. Removing pain as the fifth vital sign, implementing drug disposal programs, and returning unused medications are several measures that have been enacted (Cabrera et al., 2019). However, these measures have had little to no effect on opioid distribution and control of opioid-related deaths. In fact, there were approximately 65,000 deaths in 2016, an increase of 21% from the year prior (Jones et al., 2018). California reported six opioid related deaths per 100K people in 2018 (NIDA, 2020). Comparably, Texas reported five opioid related deaths per 100K people in 2018 (NIDA, 2020). The opioid prescription rate was 35.1 prescriptions per 100 persons in California (Huey & Apollonio, 2019) and 47.2 in Texas (NIDA, 2020) in 2018.

Due to stringent federal legislature, there has been little research deciphering the effects of medical marijuana, particularly using products available at dispensaries, and therefore the relationship between marijuana use and trends in opioid distribution and narcotic overdoses has not been fully elucidated. However, California legalized medicinal marijuana in 1996 and recreational marijuana in 2016. Recreational dispensaries opened to individuals who were at least 21 years of age in January 2018. As data demonstrating marijuana’s success as an alternative to opioids is presented (Blake et al., 2019; Boehnke et al., 2016; Kienzl et al., 2020; Lee et al., 2018; Sagy et al., 2019; Ware et al., 2015), resulting opioid trends can be analyzed. In this study, opioid distribution rates in California, where medical marijuana and recreational marijuana use was legalized, by zip code and per capita, were compared to Texas, which has not legalized recreational or medical marijuana. Texas caps the percent tetrahydrocannabinol at 0.5% and enrollment (3,519 in Dec, 2020) in their program relative to state’s population is modest (Sparber & Jones, 2021). We hypothesize that there will be a greater decrease in opioid distribution in California following the change in marijuana legislation than in Texas.

## Methods

### Procedure

The Drug Enforcement Administration’s Automation of Reports and Consolidated Orders System (ARCOS) Database is used to track controlled substances from manufacture, through distribution channels to point-of sale at the level of hospitals, pharmacies, practitioners, and teaching institutions. The data is organized in several reports summarizing drug distribution by first three-digits of the zip code, per 100K population gathered and includes summaries of purchases. Report 1, which reports drug distribution by zip code, and Report 3, which reports quarterly drug distribution by state per 100,000 population by gram were selected for analysis (*ARCOS Retail Drug Summary Report - 2011 Reporting Period*, n.d.; Ighodaro et al., 2019). Fentanyl, oxycodone, morphine, hydrocodone and codeine were chosen based on their familiarity in the general population and inclusion in prior research (Cabrerra et al., 2018; Ighodaro et al., 2019; Piper et al., 2017). ARCOS data has been previously validated by comparing results to a state Prescription Drug Monitoring Program which showed a satisfactory (r = 0.985) agreement (Piper et al., 2017).

The Center for Disease Control WONDER database was constructed to facilitate access to public health information for state and local health departments and the academic public health community (Friede et al., 1994). The mortality data was extracted from WONDER as “Drug-induced causes” set as the underlying cause of death with the Multiple Cause of Death ICD-10 codes T40.1-“Heroin” and T40.2-“Other opioids”. ICD-10 code T40.1 is limited to heroin induced deaths. ICD-10 code T40.2 excludes non-opioid narcotics, such as cocaine, opium, and LSD, and may include but is not limited to codeine, fentanyl, hydrocodone, hydromorphone, morphine, oxycodone, and tramadol. Total overdose rate includes heroin and other opioid induced deaths.

### Data Analysis

The weights of five common opioids codeine, fentanyl, hydrocodone, morphine, and oxycodone) were converted to oral Morphine Milligram Equivalents (MME) (Piper et al., 2018). Outliers were tested for using Grubbs’ Test on GraphPad Prism. Oxycodone distribution per zip code was unavailable in ARCOS for the year 2018. To account for this, oxycodone distribution in the last two quarters of 2017 was average with oxycodone distribution in the first two quarters of 2019 to represent oxycodone distribution in 2018. Heat maps and graphs were constructed using Microsoft Excel and GraphPad Prism 8. Two-tailed paired equal variance t-tests were performed using GraphPad Prism 8 software on zip code distribution data to determine statistical differences with p<0.05 considered significant.

## Results

### Opioid Distribution by Zip Code

In both 2014 and 2018, California showed much more widespread and substantial cumulative opioid distribution as seen in Fig. 1(A-B). In 2014, portions of California surrounding the Bay Area and portions of Southern California had cumulative opioid distributions as high as 1,388,000 MME and 900,000 MME, respectively. In contrast, the highest distribution in Texas was 790,000 MME in the surrounding Dallas area. Most of Texas had a distribution of 100,000 or less while most zip codes in California ranged from 150,000 to 650,000 (Fig. 1A). However, in 2018, the largest distributions in California dropped to 720,000 MME in the Bay Area and 525,000 MME in Southern California, decreases of 50 and 40 percent respectively (Fig. 1B). The surrounding Dallas area saw a decrease of 25 percent while most of Texas did not show a comparable decrease (Fig. 1C). Both California (p<0.0001) and Texas (p<0.0001) showed statistically significant decreases in opioid distribution, but California had an over two-fold difference per zip code (162,476 versus 62,808 MME) over this five-year span. This represents a −43.7% decrease in California relative to only −27.3% in Texas (Fig. 2). There was an increase of 6,000 MME in a 16-zip code area of Southern California near Glendale and Burbank. In this area, fentanyl, morphine, and oxycodone saw 1.5, 2.4, and 1.8-fold increases respectively while codeine and hydrocodone did not undergo appreciable changes (Fig. 1C).

**Figure 1.**
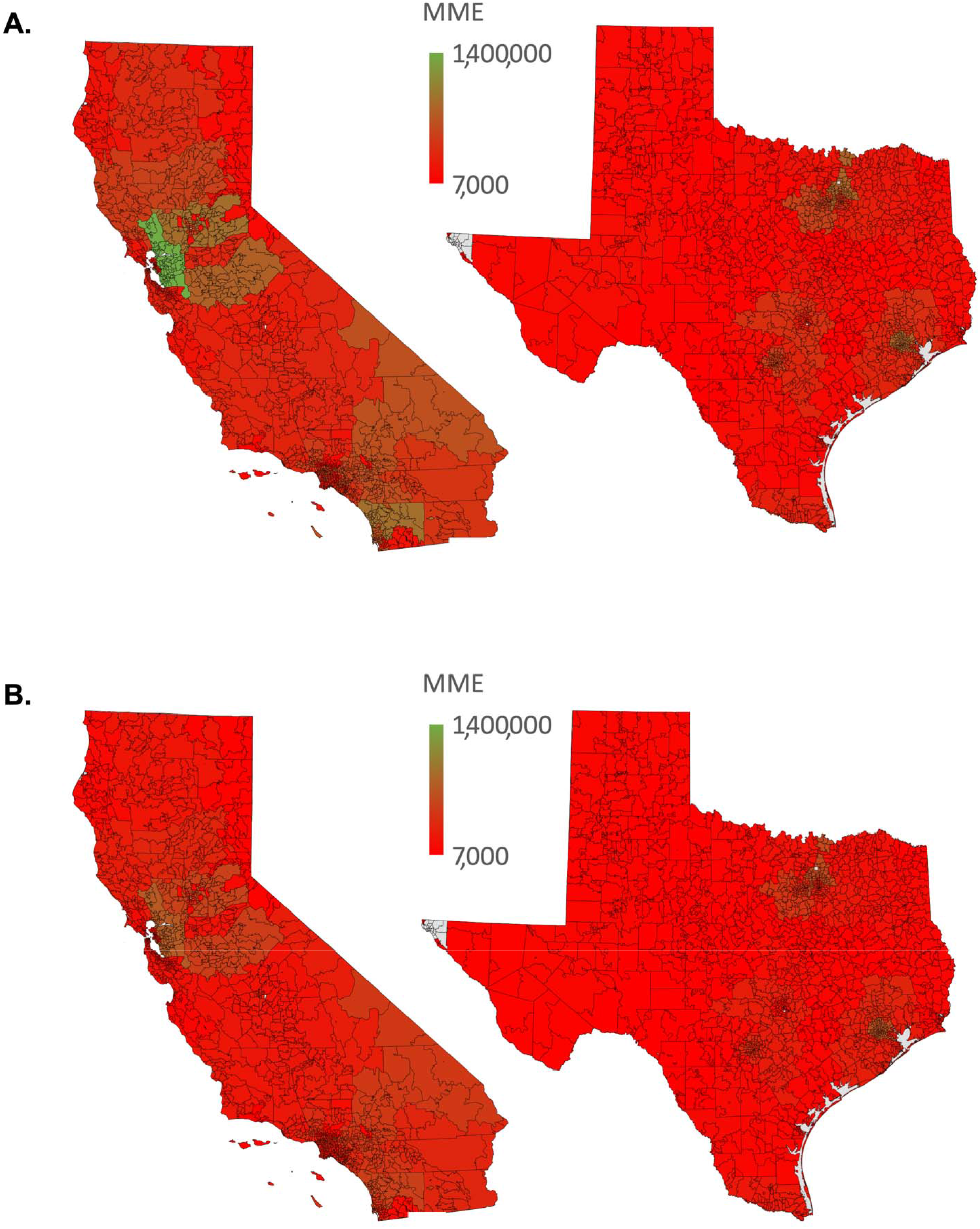
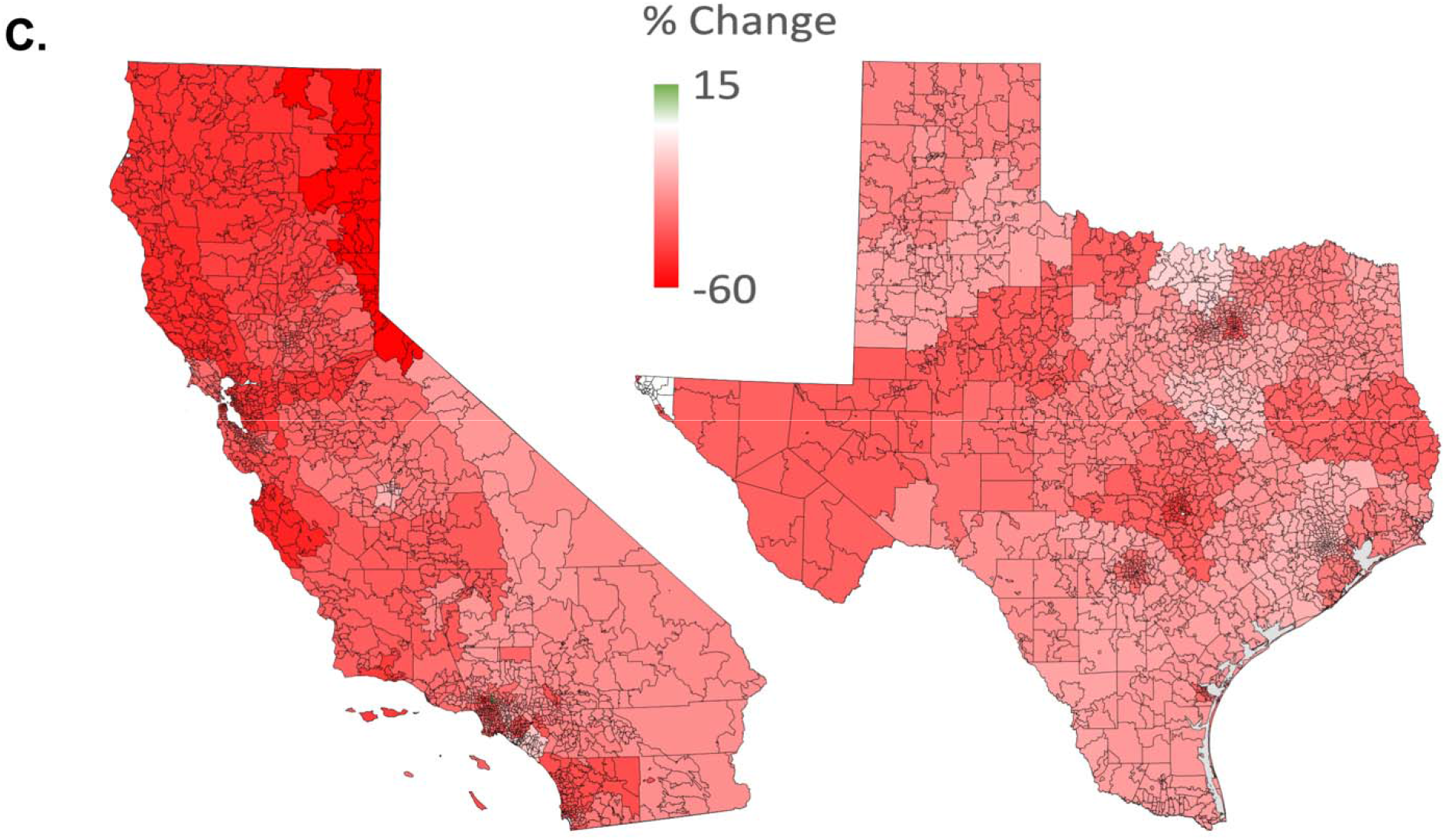
Total opioid distribution in California and Texas in Morphine Mg Equivalents (MME) by zip code as reported to the Drug Enforcement Administrations Automation of Reports and Consolidated Ordering System in 2014 **(A)** and 2018 **(B)**. Percent change from 2014 to 2018 **(C)**.

**Figure 2.**
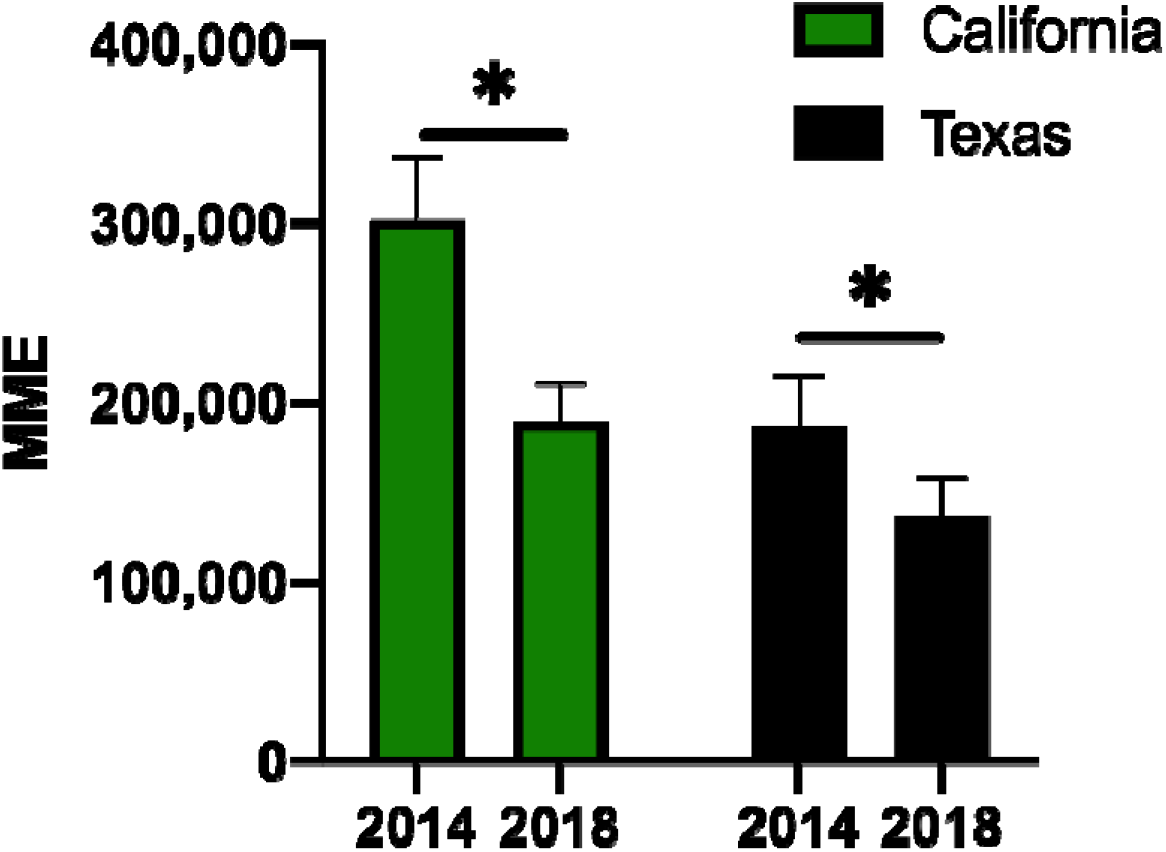
Opioid distribution (±SEM) per zip code as reported by the Drug Enforcement Administration’s Automation of Reports and Consolidated Orders System. Morphine Mg Equivalent: MME. * p < .05 versus 2014.

### Opioid Distribution Per Capita

Opioid distribution per capita decreased in both California and Texas from 2014 to 2018 (−38.9% and −26.4% respectively). The change in California was more linear for all opioids while Texas experienced more variable change (Fig. 3). Distribution of all five opioids in California decreased to a greater extent than their Texan counterparts (hydrocodone −65.4%, fentanyl −50.3%, morphine −42.7%, oxycodone −32.4%, and codeine −19.1%, Fig. 3). Individual opioid distribution changes in Texas showed much more variability with hydrocodone, fentanyl, and morphine distribution decreasing (−42.3%, −36.7%, and −27.8%), oxycodone remaining essentially unchanged (−0.5%), and codeine distribution increasing 150.1% (Fig. 3).

**Figure 3.**
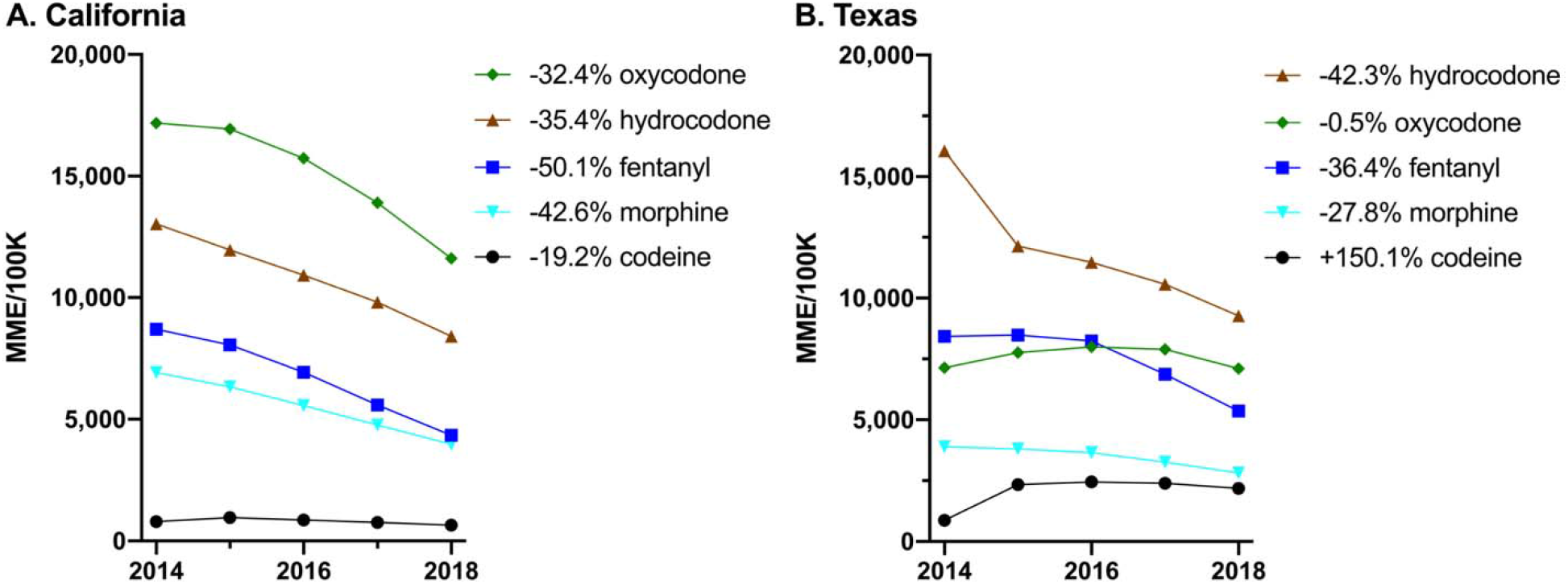
Per capita distribution of selected opioids from 2014 to 2018 as reported to the Drug Enforcement Administration’s Automation of Reports and Consolidated Orders System in California **(A)** and Texas **(B)**. The percent change from 2014 to 2018 is shown in the key.

### Overdose rates

Overall, opioid related deaths increased in California (11.6%) and Texas (272.7%) between 1999 and 2019. California showed a decline in deaths from non-heroin opioids between 2014-2019 (Fig. 4A) while Texas had an increase in deaths from 2014-2017 and then experienced a decline in 2018 (Fig. 4B). Heroin related deaths have increased continuously in California (71.4%) and Texas (400%) between 1999 and 2019.

**Figure 4.**
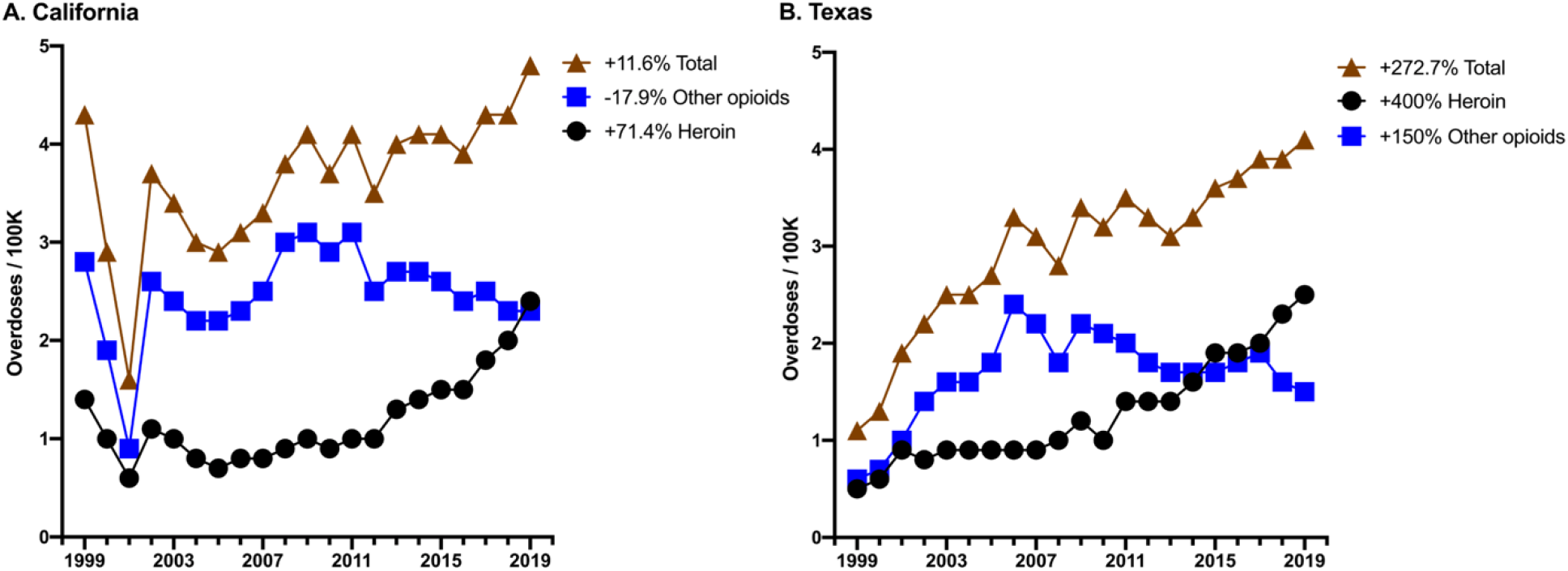
Opioid overdose rates as reported by the CDC’s Wide Ranging Online Data for Epidemiologic Research (WONDER) from 1999 to 2019 in California **(A)** and Texas **(B)**. The percent change from 1999 to 2019 is listed in the key.

## Discussion

The observed trends in opioid distribution by zip code and per capita in California and Texas support recreational marijuana legalization as a practical measure to help reduce prescription opioid use. The five years analyzed (2014 – 2018) show a greater decrease in California than in Texas (Fig. 2). Opioid distribution per 100K people in California decreased 38.9% from 2014-2018 while distribution in Texas decreased by only 26.4% (Fig. 3). Similarly, heat map analysis (Fig. 1(A-B) illustrates a greater decrease in opioid distribution over large areas of California compared to Texas from 2014 to 2018. These findings are consistent with investigations showing fewer opioid prescriptions written in states where medical marijuana laws are passed (Bradford & Bradford, 2016). A similar study showed a significant correlation between increased marijuana use as a treatment for chronic pain and reduced opioid use (Boehnke et al., 2016), suggesting a potential public health benefit of replacing opioids with cannabis.

The drop in hydrocodone and increase in codeine and oxycodone distribution per capita seen in Texas (Fig. 3) is consistent with previous data showing that oxycodone and codeine increased by weight distribution from 2014 to 2017 specifically in Texas (NIDA, 2020). The reclassification of hydrocodone in 2014 from a schedule III to a schedule II may be a factor behind this trend (Bohnert et al., 2018; NIDA, 2020). Increased restrictions surrounding hydrocodone may have created an environment for the distribution of oxycodone and codeine to rise as there were fewer powerful analgesic options available, such as cannabis products.

The decreasing trends observed in California (Fig.3) may be explained by current approaches to treating pain and the reduction in opioid use as a result of medicinal and recreational marijuana legalization. Three-fifths (62%) of women surveyed in California and Colorado with gynecological malignancies stated their interest in non-prescription cannabis and 35.6% were interested in using cannabis under the guidance of physicians. One-quarter (26.7%) of these patients reported already using non-prescription cannabis products to manage various cancer related symptoms (Blake et al., 2019).

California saw a decrease in overdose deaths after the legalization of marijuana in 2016 and 2018 (Fig.4), whereas Texas, which has not legalized marijuana, experienced a rise in deaths by overdose in 2016 and 2017 however a gradual decrease in 2018 was also shown. Results from our data could be accounted for by the implementation of laws across the U.S. designed to mitigate overdoses by limiting opioid prescriptions and institutionalizing Prescription Drug Monitoring Programs introduced in 2017 (McGinty et al., 2018). Prior research has found that any association between medical marijuana and opioid overdoses is complicated (Bachhuber et al., 2014; Kaufman et al., 2021; Shover et al., 2019), but further studies will help describe the raltionship between changing marijuana legislature and opioid overdoses.

Although harm reduction policies and medication-assisted treatments for opioid dependence have increased in availability, (e.g. sterile syringe access programs and methadone) (Bohnert et al., 2018), alternative pain management strategies must be employed to decrease access and use of opioids to lower addiction and overdose rates. States that enacted medical cannabis laws had a 24.8% reduction in opioid overdose mortality which further decreased in the years following their implementation (Bachhuber et al., 2014). Evidence supports the promotion of safer and less addictive alternatives for analgesic relief, and implementation of laws that could reduce the misuse and abuse of opioids.

There were a variety of limitations to our study. Limited research on recreational and non-prescription marijuana for analgesic effects makes it difficult to make accurate analysis and inferences. Most findings on cannabis and chronic pain refer to medical cannabis, but patients have reported using recreational and non-prescription cannabis for medical benefits (Blake et al., 2019). Surveys taken to assess the use of recreational and non-prescription marijuana from dispensaries would help differentiate between recreational and medicinal marijuana use. The data extracted from ARCOS presented a further limitation as the census data used by the Drug Enforcement Administration is from 2010. Including more control states and utilizing 2020 census data, when available, might be beneficial in future studies (Andreae et al., 2016). In addition, ARCOS does not account for mail-order pharmacies and internet pharmacies that ship different prescription opioids across state lines, including the five opioids used in this study (NIDA, 2020). Lastly, statistics gathered from CDC WONDER database do not provide specific details on overdose outcomes and includes overdoses from all opioids, not just the five previously discussed. Overdose data from WONDER may also be underestimated and accuracy varies from state to state as there is no standardized national reporting practice (NIDA, 2020; NIDA, 2020; Kaufman et al., 2021). However, both California and Texas were rated by the CDC as in the same “less than good” category for death determination quality (Kaufman et al., 2021).

## Conclusion

Prescription opioid distribution in California showed a greater decrease compared to Texas (43.7% and 27.3% respectively). These findings suggest a relationship between California’s marijuana legislation and the decreased opioid distribution when compared to a state that has legislation prohibiting medicinal or recreational marijuana use. As more states increase access to cannabis, trends in opioid distribution and use, as well as opioid overdoses, must be further analyzed to promote legislature to help combat the opioid epidemic.

## Data Availability

Data from this manuscript can be obtained online from CDC WONDER online database and the Automated Reports and Consolidated Ordering System (ARCOS)

https://www.deadiversion.usdoj.gov/arcos/retail_drug_summary/

https://wonder.cdc.gov/

## Author Contributions

Michelle N Anyaehie conceived and designed the study, performed literature search and review, authored drafts of the paper, analyzed the data and approved the final manuscript.

Christian Pardo conceived and designed the study, preformed literature search and review, analyzed the data, prepared figures, authored drafts of the paper, and approved the final manuscript.

Elijah J Johnson conceived and designed the study, preformed literature search and review, authored drafts of the paper, and approved the final manuscript.

Chucks Anachebe authored drafts of the paper and approved the final manuscript.

Dr. Brian Piper authored drafts of the paper, analyzed data, and approved the final manuscript.

## Acknowledgments

We would like to thank Elizabeth Kuchinski, MPH and Dan Kaufman, MBS at Geisinger Commonwealth School of Medicine for their guidance and feedback throughout this study.

## Supplemental Figures

**Figure S1.**
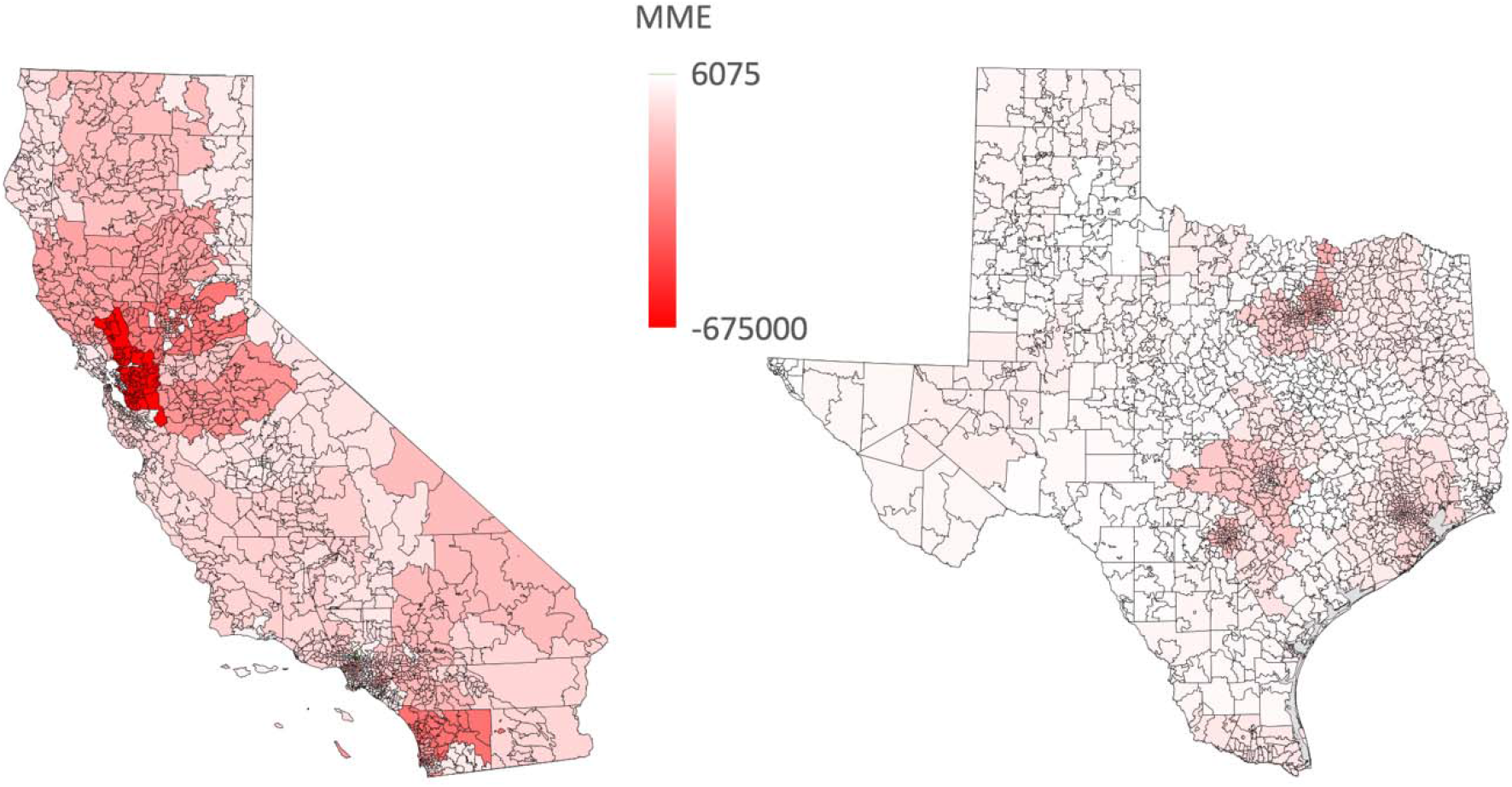
Cumulative change in opioid distribution as reported to the Drug Enforcement Administration’s Automation of Reports and Consolidated Orders System in California and Texas from 2014 to 2018.

**Figure S2.**
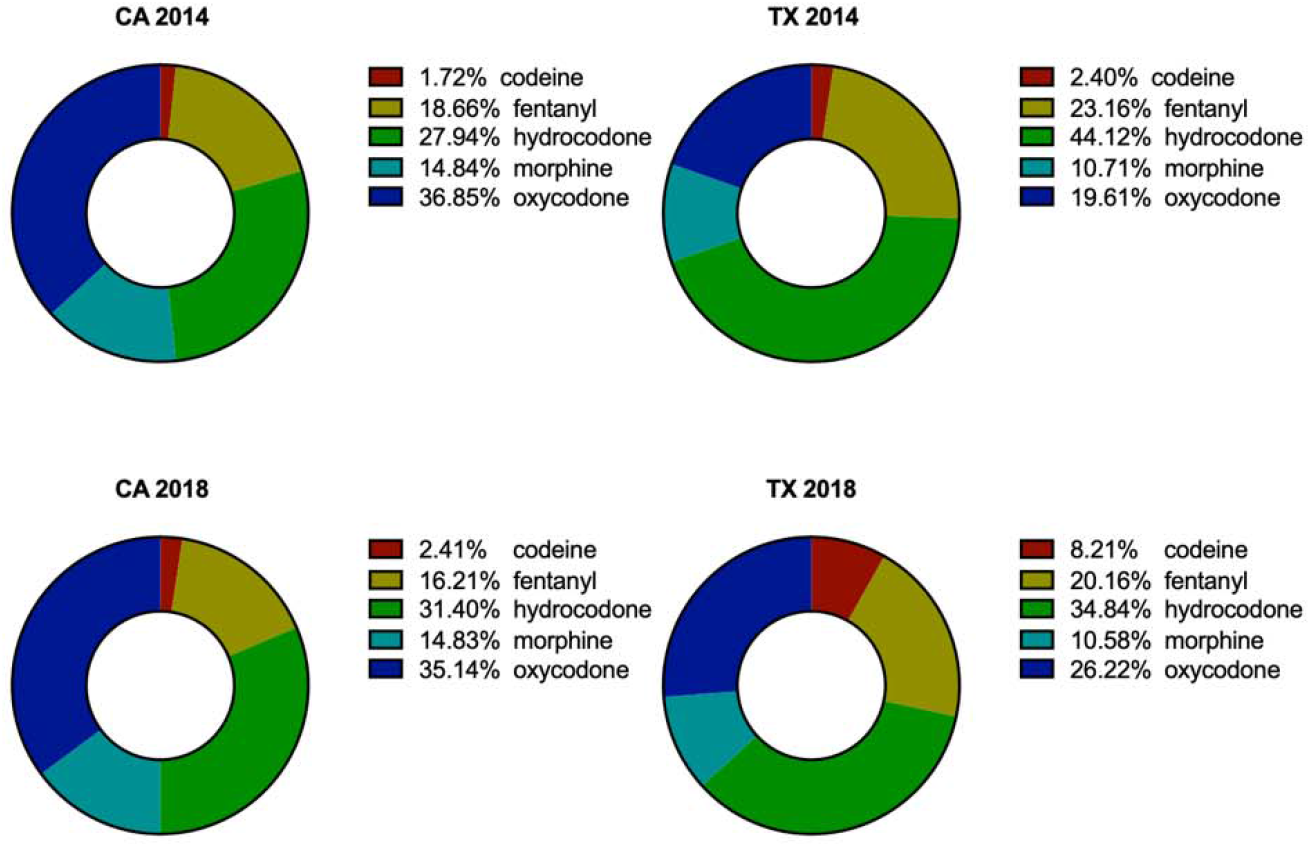
Opioid composition as reported to the Drug Enforcement Administration’s Automation of Reports and Consolidated Orders System per state (California : CA, Texas: TX) in 2014 (top) and 2018 (bottom).

